# Mepolizumab effectiveness in patients with severe eosinophilic asthma and co-presence of bronchiectasis: a real-world retrospective pilot study

**DOI:** 10.1101/2020.11.04.20226266

**Authors:** Claudia Crimi, Raffaele Campisi, Santi Nolasco, Giulia Cacopardo, Rossella Intravaia, Morena Porto, Pietro Impellizzeri, Corrado Pelaia, Nunzio Crimi

## Abstract

**Background:** The association of bronchiectasis (BE) in patients with severe eosinophilic asthma (SEA) is quite frequent. Mepolizumab is a well-recognized treatment for SEA; we aim to evaluate its effectiveness in SEA patients with and without BE in real-life.

**Methods:** We performed a single-center retrospective pilot study, including patients with SEA treated with mepolizumab for one year. Asthma control test (ACT), lung function, annual exacerbations rate, oral corticosteroid dosage, FeNO, chronic mucous secretions, blood and sputum eosinophils were recorded at baseline and after 6 and 12 months.

**Results:** we included 32 patients (mean age: 52.3±10, 59% female). 50% showed co-presence of bronchiectasis, (SEA+BE). Significant improvements were found in ACT [(13.8±4.6 to 20.7±4.1, p=0.0009) and (13±4.8 to 20.7±4.6, p=0.0003)], annual exacerbations rate [from 7 (4-12) to 0 (0.00-0.75) and from 8 (4-12) to 0 (0-1), p<0.0001], and blood eosinophils count [748 cells/μL (400-1250) vs. 84 cells/μL (52.5-100), and from 691 cells/μL (405-798) vs. 60 cells/μL (41-105), p<0.0001] in SEA and SEA+BE group respectively, already after 6 months of treatment. A reduction in daily oral corticosteroids intake at 12 months was shown [from 15 mg (0-25) to 0 mg (0-0), p=0.003 and from 8.8 mg (0-25) to 0 mg (0-0) (p=0.01)] in both SEA and SEA+BE, respectively. Similar results were found, comparing SEA+BE patients based on the severity of bronchiectasis.

**Conclusions:** Mepolizumab effectively improves asthma symptoms control, reducing annual exacerbations and corticosteroid intake in all patients with SEA, even in the subgroup with coexisting bronchiectasis, independently of their severity.

**Highlights:**

- Mepolizumab is effective in the treatment of severe eosinophilic asthma (SEA) in clinical trials and real-life studies.
- Bronchiectasis is a frequent comorbidity in patients with severe eosinophilic asthma.
- Mepolizumab proved to be effective in improving asthma symptoms control, mucus hypersecretion, lung function, and reducing sputum and blood eosinophils, corticosteroids dependency and annual exacerbations both in severe eosinophilic asthma patients with or without co-presence of bronchiectasis.
- Mepolizumab showed to be effective in patients with asthma and co-presence of bronchiectasis, regardless of BE severity.
- Assessing the co-presence and severity of bronchiectasis may help clinicians select the right biologic for a prompt treatment-specific effect.

## INTRODUCTION

Severe refractory asthma is a heterogeneous disease with a high burden on the health care system [1-2]. Several comorbidities and risk factors [3] are frequently associated with severe asthma, identifying different phenotypes [4], and their recognition and treatment are crucial to improving asthma control and outcomes [5].

The coexistence of asthma and bronchiectasis (BE) represents an emerging phenotype [6] associated with a worse prognosis of the disease [7]. The prevalence of BE is higher in patients with severe eosinophilic asthma (SEA), ranging from 24 to 40% [5-9], and it is characterized by a late-onset disease, older age, a non-reversible airflow obstruction, rhinosinusitis, chronic mucus expectoration and frequent or severe exacerbations [10-11].

Mepolizumab, a recently released IL-5 antagonist, has shown to be an effective treatment for SEA [12], reducing exacerbations, the number of blood and sputum eosinophils, and improving pulmonary function and, therefore, asthma control [13]. Moreover, the long-term eosinophils-mediated inflammatory damage [14-15], sustained by IL-5, induces tissue changes and airway remodeling, playing a crucial role in the pathogenesis of bronchiectasis [14-16].

We hypothesized that mepolizumab would effectively improve asthma symptoms control and reduce exacerbations in a cohort of SEA with or without associated BE. We aim to evaluate the effectiveness and clinical outcomes of mepolizumab treatment in SEA patients with and without coexistence of BE and to verify its efficacy in patients with SEA+BE, according to BE severity.

## METHODS

### Study design and patient population

We performed a single-center, retrospective observational study based on medical records review of patients with SEA, who were referred to the Respiratory Medicine Unit - AOU “Policlinico-Vittorio Emanuele”, Catania - Italy, between December 2018 and January 2020. This study adhered to the Declaration of Helsinki and received approval from the Ethics Committee “Catania 1” at Policlinico Hospital (Protocol Number 33/2020/PO).

We included all adult outpatients diagnosed with SEA according to the European Respiratory Society/American Thoracic Society (ERS/ATS) criteria [1] and Global Initiative for Asthma (GINA) 2019 report [17], requiring GINA step 4-5 treatment (high-dose inhaled corticosteroids (ICS), long-acting β-agonists (LABAs) and long-acting muscarinic receptor antagonists (LAMAs), or systemic corticosteroids ≥ 50% of the previous year to remain controlled or that remains “uncontrolled” despite this treatment) [17], prescribed with mepolizumab 100 mg subcutaneously every 4 weeks, with a continuous follow-up of at least one year.

### Data collection and assessment

An established database of pertinent variables was accessed for data analysis. Socio-demographic characteristics (age, sex, BMI) and baseline asthmatic profiles information [age at onset, skin prick test to evaluate sensitization to common aeroallergen, pulmonary function test, blood eosinophils count, IgE, fractional exhaled nitric oxide (FeNO) measurement, Asthma Control Test (ACT) score [18], sputum cytology analysis, asthma exacerbation/hospitalization rate and maintenances therapies, daily corticosteroid intake (prednisone mg/day)], were retrieved from the database as well as the presence of comorbidities [nasal polyps, rhinosinusitis, chronic mucus hypersecretion, gastro-esophageal reflux (GERD), obesity, and bronchiectasis].

All these information were collected at baseline (T0) before mepolizumab initiation, and after 6 (T6) and 12 (T12) months of biologic treatment initiation.

Asthma exacerbations were defined as disease worsening, requiring an emergency department visit, hospitalization, and/or the use of OCS for ≥48 h or an increase of ≥50% in daily OCS dose [19].

### Pulmonary function

Pulmonary function tests were performed according to ERS/ATS guidelines [20]. Forced Vital Capacity (FVC) and Forced Expiratory Volume in 1 second (FEV_1_) were measured using a spirometer (Sensormedics, Milan, Italy). The best value of three consecutive maneuvers was expressed as the percentage of the normal. After the baseline assessment, spirometry was repeated 15 minutes after salbutamol administration (400 mcg). Reversibility of airway obstruction was expressed as a change of at least 12% of the FEV_1_ from baseline.

### Asthma Control Test

Asthma control was assessed at baseline (T0) and after 6 (T6) and 12 (T12) months of treatment with mepolizumab using the ACT [18]. An overall score of at least 20 refers to well-controlled asthma, whereas a score ≤19 reflects poor control.

### FeNO Measurements

The fraction of exhaled nitric oxide was performed according to ERS/ATS guidelines [21] and was measured using the Niox ® device. FeNO levels were considered normal if FeNO was less than 25 parts per billion (ppb) and elevated if FeNO was higher than 50 ppb, as recommended [22].

### Induced sputum

Sputum was induced through the inhalation of hypertonic saline solution (4.5%) using an ultrasonic nebulizer (DeVilbiss 65; DeVilbiss Corporation, Somerset, PA, USA) [23]; sputum samples were then processed for cytological analysis.

### Gastro-esophageal reflux disease

Gastro-esophageal reflux (GERD) diagnosis was based on patients’ symptoms (heartburn and acid regurgitation) and accurate clinical history, such as extensive use of NSAIDs, as suggested by societal guidelines [24].

### Chronic Rhinosinusitis with Nasal Polyps

The diagnosis of chronic rhinosinusitis with nasal polyps (CRSwNP) was formulated based on symptoms (nasal congestion, nasal discharge, facial pain/pressure, reduction/loss of smell for a minimum of 12 weeks) and objective confirmation of the presence of polyps with sinus computed tomography (CT) scan or nasal endoscopy, according to the European position paper on rhinosinusitis and nasal polyps (EPOS) 2020 [25].

### Bronchiectasis diagnosis and severity

All the patients underwent high resolution computed tomography (HRCT) of the lung, as a routine exam performed in the context of severe asthma investigations in our center. The diagnosis of bronchiectasis was made based on the identification of a broncho-arterial ratio greater than 1.5 [26]. All HRCT were reported by radiologists with specific expertise in high-resolution imaging. An expert thoracic radiologist and a pulmonologist subsequently performed an independent review to confirm and score disease severity.

Bronchiectasis Severity Index (BSI) score [27] was calculated in patients affected by bronchiectasis, combining body mass index (BMI), FEV_1_% predicted, hospital admission in the past two years, number of exacerbations in the previous year, Modified Medical Research Council (MMRC) breathlessness score, radiological severity (≥ 3 lobes involved or cystic changes), presence of pseudomonas and chronic bacteria colonization [28]. Microbiology testing was performed on patients’ spontaneous early morning sputum samples. According to the BSI overall score (range from 0 to 26), bronchiectasis was defined as mild (BSI = 0–4 points), moderate (BSI = 5–8 points), or severe (BSI ≥9 points) [29].

### Chronic Mucus Hypersecretion

Symptoms of chronic mucus hypersecretion (CMH), defined as cough and sputum production on most days for at least three months a year for at least two consecutive years, were recorded [30].

### Statistical analysis

Categorical variables are stated as number (n) and percentage (%); continuous variables are expressed as mean ± standard deviation (SD) or median and interquartile range (IQR), as appropriate. Patients were divided into two groups: SEA alone and SEA+BE, to allow for comparisons and outcomes. We further classified SEA+BE patients into two groups based on bronchiectasis severity: 1) mild BSI group and 2) moderate-to-severe BSI group.

The normality of data distribution was checked using the Shapiro-Wilk test and the Kolmogorov-Smirnov test. Fisher exact test was used for comparisons of categorical variables. Linear regression and Spearman’s rank Correlation Coefficient were used to examine the relationship between variables. ANOVA for repeated measures was used for comparisons among T0 (baseline), T6 (after 6 months of mepolizumab treatment) and T12 (after 12 months of mepolizumab treatment) for continuous variables, followed by Bonferroni’s correction. Friedman or Kruskal-Wallis tests were used for comparisons among T0, T6, and T12 of non-parametric variables, followed by Dunn post-hoc. In case of direct comparison between T0 and T12, paired t-test or Wilcoxon tests were used for continuous and non-normally distributed variables. Unpaired t-test or Mann-Whitney were used for comparisons between groups, as appropriate. Statistical analysis was performed using Prism version 8.2.1 (GraphPad Software Inc). A p value < 0.05 was considered statistically significant.

## RESULTS

### Patient demographics and clinical characteristics

A total of 69 patients were referred for evaluation of SEA. Thirty-two patients treated with mepolizumab and followed-up for 12 months were included in the study. The mean age of patients was 52.3±10 years old. There was a female predominance (19/32 patients, 59%). Sixteen out of 32 patients (50%) showed co-presence of bronchiectasis at chest HRCT and were identified as SEA+BE group. Baseline characteristics of patients and comparisons between SEA alone and SEA+BE are reported in Table 1. No statistically significant differences in terms of ACT, FEV_1_%, and blood eosinophil levels were noted between the groups. Annual exacerbations and daily OCS intake were more pronounced in the SEA+BE group than in SEA alone, but this difference did not reach a statistical significance. A greater proportion of patients suffered from CMH in the SEA+BE group than in the SEA alone [14 (87.5) vs. 5 (31.24), respectively, p=0.0032].

**Table 1.** Patients’ baseline demographic and clinical characteristics. Abbreviations: SEA, Severe Eosinophilic Asthma; BE, Bronchiectasis; BMI, body mass index; CRSwNP, Chronic Rhinosinusitis with Nasal Polyps; GERD, Gastroesophageal reflux disease; ICS-LABA, Inhaled Corticosteroids - Long-Acting Beta-Agonist; LAMA, Long-Acting Muscarinic Antagonist; CMH, Chronic Mucus Hypersecretion; OCS, oral corticosteroids (Prednisone); ACT, Asthma Control Test; FEV1, Forced expiratory volume in the 1st second; IgE, Immunoglobulin-E; FeNO, Fractional Exhaled Nitric Oxide; SD, standard deviation; IQR, interquartile range; ppb, parts per billion.

|  | Total Population<br>(n=32) | SEA<br>(n=16) | SEA+BE<br>(n=16) | p |
| --- | --- | --- | --- | --- |
| Age, years, mean (SD) | 52.3 (10) | 53.2 (11.9) | 51.3 (7.9) | 0.60 |
| Male, n (%) | 13 (40.6) | 5 (31.3) | 8 (50) | 0.47 |
| BMI, mean (SD) | 26.7 (5.2) | 28.3 (5.6) | 25.1 (4.1) | 0.08 |
| Duration of asthma, years, mean (SD) | 19.5 (12.5) | 17.8 (12.8) | 21.1 (12.4) | 0.46 |
| Positive Skin Prick Test, n (%) | 23 (71.9) | 12 (75) | 11 (68.8) | >0.99 |
| CRSwNP, n (%) | 24 (75) | 12 (75) | 12 (75) | >0.99 |
| GERD, n (%) | 11 (34.4) | 8 (50) | 3 (18.8) | 0.13 |
| Asthma exacerbations/year, median (IQR) | 7(4-12) | 7 (4-12) | 8 (4-12) | 0.88 |
| ICS-LABA, n (%) | 32 (100) | 16 (100) | 16 (100) | >0.99 |
| LAMA, n (%) | 20 (62.5) | 9 (56.3) | 11 (68.8) | 0.71 |
| Patients with CMH, n (%) | 19 (59.37) | 5 (31.25) | 14 (87.5) | **<0.01** |
| Patients on OCS, n, (%) | 21 (65.6) | 11 (68.8) | 10 (62.5) | >0.99 |
| OCS, mg/die, median (IQR) | 8.8 (0-25) | 8.8 (0-25) | 15 (0-25) | 0.68 |
| ACT, mean (SD) | 13.4 (4.7) | 13.8 (4.6) | 13 (4.8) | 0.63 |
| FEV<sub>1</sub>, %, mean (SD) | 75.1 (22.6) | 76.6 (20.9) | 73.7 (24.8) | 0.72 |
| Blood Eosinophils, cells/μL median (IQR) | 795 (427-1308) | 748 (400-1250) | 691 (405-798) | 0.46 |
| IgE, UI/ml, median (IQR) | 213 (92-414) | 260 (92-505) | 177 (95-329) | 0.31 |
| FeNO, ppb, mean (SD) | 49.1 (42.8) | 49.8 (39.4) | 48.3 (47.2) | 0.78 |
| Sputum eosinophils, %, median (IQR) | 22.5 (16.3-49) | 20.5 (17.3-42.3) | 26.5 (10.8-62.8) | 0.90 |
| Sputum neutrophils, %, median (IQR) | 17 (8.00-27.8) | 14.5 (7-20.8) | 24.5 (8.25-48.3) | 0.06 |

### Indices of Mepolizumab effectiveness

Changes in indices of mepolizumab effectiveness are shown in Figure 1. The ACT score significantly improved from baseline, already after six months of mepolizumab treatment (T6) in both SEA (13.8±4.6 to 20.7±4.1, p=0.0009) and SEA+BE (13±4.8 to 20.7±4.6, p=0.0003) group, respectively. This trend of ACT score improvement was maintained during the entire study follow up, at T12, reaching a mean ACT score value of 21.9±3.1 (p<0.0001) in the SAE alone group, Figure 1 panel A. Asthma exacerbations/year significantly lowered overall, going from 8 (4-12) to 0 (0-1) in SEA+BE and from 7 (4-12) to 0 (0.00-0.75) in SEA group, respectively (p<0.0001), Figure 1 panel B. Daily OCS dose significantly decreased at T12, changing by a median of 15 mg (0-25) to 0 mg (0-0) in SEA+BE group (p=0.003) and from 8.8 mg (0-25) to 0 mg (0-0) in SEA alone group (p=0.01), as shown in Figure 1 panel C. A significant change in mean predicted FEV_1_ was observed only in the SEA+BE group, from 73.7%±24.8% at T0 to 85.9%±18.3% at T6 (p=0.02), as shown in Figure 1 panel D. No differences in FeNO values after treatment were noted in both groups. Chronic Mucus Hypersecretion significantly decreased in the SEA+BE group after treatment (p=0.0011), as shown in Figure 1panel E. Blood eosinophils count was significantly reduced from baseline to T6 [a median of 748 cells/μL (400-1250) vs. 84 cells/μL (52.5-100), (p<0.0001)] in SEA and SEA+BE from 691 cells/μL (405-798) to 60 cells/μL (41-105). In SEA+BE, blood eosinophils further decreased to 49 cells/μL (32.5-90), (p<0.0001), with a statistically significant difference in comparison to SEA at T12 (p=0.04), Figure 1 panel F. Sputum eosinophil count reduced from 26.5% (10.8-62.8) to 12.5% (5-23) (p<0.0001) in SEA+BE group and from 20.5% (17.3-42.3) to 9% (7-10) at (p<0.0001) in SEA group, Figure 2 panel A. Sputum neutrophils decreased from 24.5% (8.3-48.3) to 14.50% (4-29.8) in SEA+BE group (p=0.012), Figure 2 panel B. Indices of mepolizumab effectiveness are reported in Table 2.

**Table 2.** Indices of Mepolizumab effectiveness in severe asthma with (SEA+BE) or without Bronchiectasis (SEA). *p<0.05 T0 vs. T6/T12 *Abbreviations: SEA, Severe Eosinophilic Asthma; BE, Bronchiectasis; BMI, body mass index; CRSwNP, Chronic Rhinosinusitis with Nasal Polyps; GERD, Gastroesophageal reflux disease; ICS-LABA, Inhaled Corticosteroids - Long-Acting Beta-Agonist; LAMA, Long-Acting Muscarinic Antagonist; CMH, Chronic Mucus Hypersecretion; OCS, oral corticosteroids (Prednisone); ACT, Asthma Control Test; FEV*_*1*_, *Forced expiratory volume in the 1st second; IgE, Immunoglobulin-E; FeNO, Fractional Exhaled Nitric Oxide; SD, standard deviation; IQR, interquartile range; ppb, parts per billion, T0, baseline; T6, follow up at 6 months; T12, follow-up at 12 months*.

|  | SEA | SEA+BE | p | SEA | SEA+BE | p | SEA | SEA+BE | p |
| --- | --- | --- | --- | --- | --- | --- | --- | --- | --- |
| ACT, mean (SD) | 13.8 (4.7) | 13 (4.8) | 0.63 | 20.7 (4.1) * | 20.7 (4.6) * | 0.89 | 21.8 (3.1) * | 20.7 (5.5) * | 0.67 |
| Asthma exacerbations/year, median (IQR) | 7 (4-12) | 8 (4-12) | 0.88 | ---- | ---- |  | 0 (0-0.8) * | 0 (0-1) * | 0.75 |
| OCS, mg/die, median (IQR) | 8.8 (0-25) | 15 (0-25) | 0.68 | 0 (0-5) | 0 (0-5) | 0.99 | 0 (0-0) * | 0 (0-0) * | 0.99 |
| FEV<sub>1</sub>, %, mean (SD) | 76.6 (20.9) | 73.7 (24.8) | 0.72 | 83.9 (21.8) | 85.9 (18.3) * | 0.72 | 83.1 (20.2) | 82.4 (21.8) | 0.87 |
| CMH, n (%) | 5 (31.25) | 14 (87.5) | <0.01 | ---- | ---- |  | 3 (18.75) | 4 (25.0) * | 0.68 |
| Blood Eosinophils, cells/μL, median (IQR) | 748 (400-1250) | 691 (405-798) | 0.46 | 84 (52.5-100) * | 60 (41-105) * | 0.32 | 90 (60-117) * | 49 (32.5-90) * | 0.04 |
| FeNO, ppb, mean (SD) | 49.9 (±39.4) | 48.3 (47.2) | 0.78 | 60.1 (46.5) | 59.8 (51.7) | 0.80 | 42.9 (23.3) | 60.2 (47.7) | 0.10 |
| Sputum eosinophils, %, median (IQR) | 20.5 (17.3-42.3) | 26.5 (10.8-62.8) | 0.90 | ---- | ---- |  | 9 (7-10) * | 12.5 (5-23) * | 0.41 |
| Sputum neutrophils, %, median (IQR) | 14.5 (7-20.8) | 24.5 (8.3-48.3) | 0.06 | ---- | ---- |  | 9.5 (5.3-17.5) | 14.5 (4-29.8) * | 0.33 |

**Figure 1.**
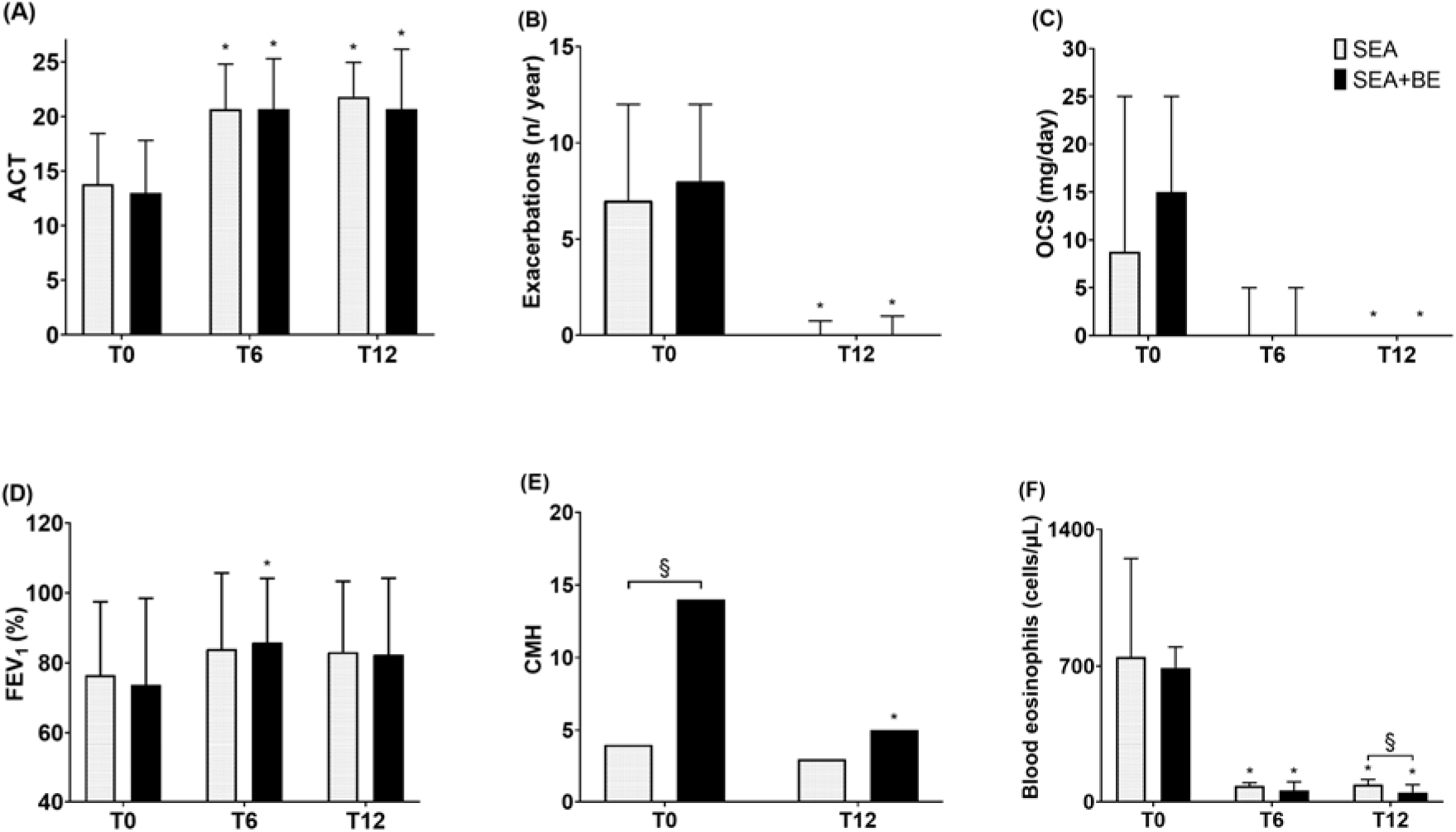
Effects of mepolizumab on SEA and SEA+BE groups, on ACT score (panel A), annual asthma exacerbations rate (panel B), OCS intake (panel C), FEV_1_ (panel D), blood eosinophils (panel F). ACT score and FEV_1_ values are expressed as mean (SD). All other parameters are expressed as median values (IQR). *p<0.05 T0 vs. T6/T12,§ p<0.05 SEA vs. SEA+BE. *Abbreviations: SEA, Severe Eosinophilic Asthma; BE, Bronchiectasis; ACT, Asthma Control Test; OCS, oral corticosteroids (Prednisone); FEV*_*1*_, *Forced expiratory volume in the 1st second; CMH, Chronic Mucus Hypersecretion*.

**Figure 2.**
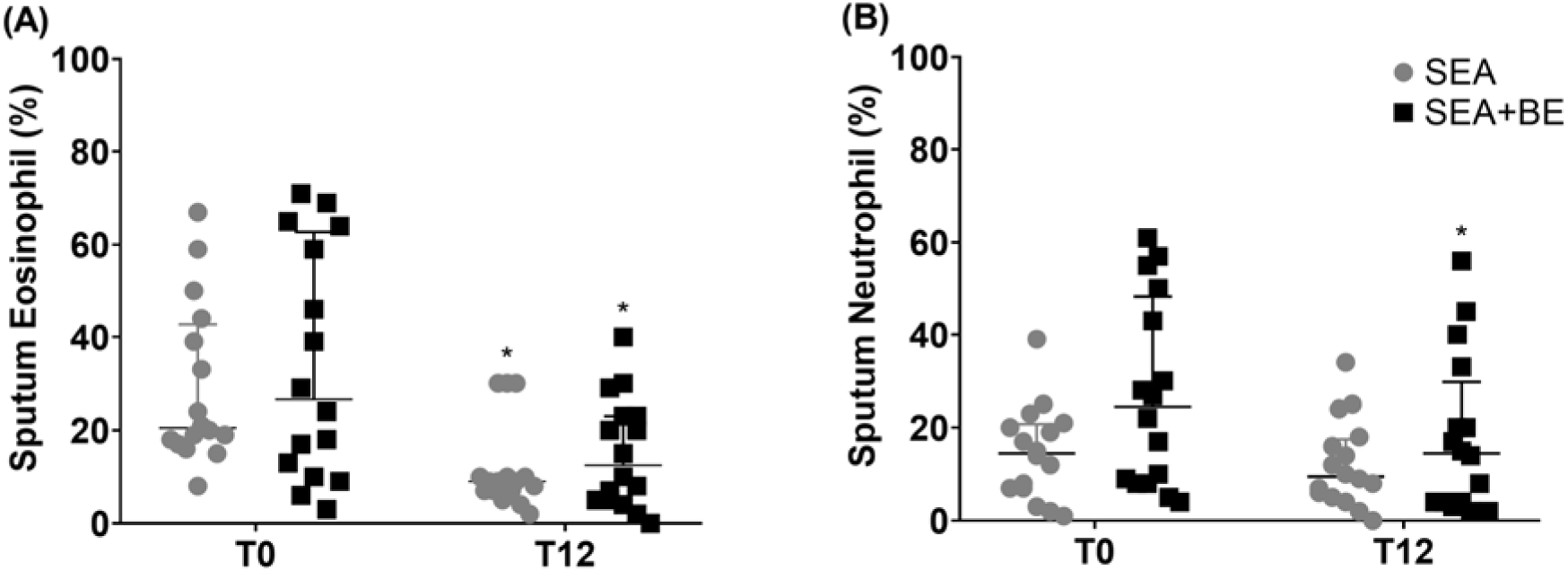
Effects of mepolizumab on sputum eosinophils percentage (%), panel A and sputum neutrophil percentage (%), panel B. *p<0.05 T0 vs. T12 *Abbreviations: SEA, Severe Eosinophilic Asthma; BE, Bronchiectasis; T0, baseline; T12, follow-up at 12 months*.

### Mepolizumab effectiveness according to BSI

Taking into account only patients with SAE+BE, we calculated BSI in 16 patients (mean 5.5±2.3). We found a significant inverse linear relationship at T0 between BSI and ACT score (Figure 3A; r = −0.7381, p=0.0016) and between BSI and FEV_1_ % (Figure 3B; r = −0.5962, p=0.0165), respectively. No significant correlation was found between BSI and duration of asthma (r = 0.14, p=0.61), asthma exacerbation (r=0.07, p=0.8), prednisone daily dose (r=0.06, p=0.83), eosinophil count in sputum (r=0.4, p=0.12), neutrophil count in sputum (r=-0.15, p=0.6), peripheral blood eosinophil count (r= 0.04, p=0.9), IgE (r= −0.43, p=0.0958), and FeNO (r = 0.07, p=0.8). Patients were further divided according to BSI score into two groups: 1) mild BSI (0–4 points) group (7 patients, 43.7%); and 2) moderate-to-severe BSI group [7 patients, (43.7%) with intermediate BSI (5–8 points), and 2 patients (12.5%) with high BSI (≥9 points)], as shown in Table 3.

**Table 3.** Baseline demographic and clinical characteristics of Mild BSI and Moderate-to-Severe BSI patients. Abbreviations: SEA, Severe Eosinophilic Asthma; BE, Bronchiectasis; BMI, body mass index; CRSwNP, Chronic Rhinosinusitis with Nasal Polyps; GERD, Gastroesophageal reflux disease; ICS-LABA, Inhaled Corticosteroids - Long-Acting Beta-Agonist; LAMA, Long-Acting Muscarinic Antagonist; CMH, Chronic Mucus Hypersecretion; OCS, oral corticosteroids (Prednisone); ACT, Asthma Control Test; FEV1, Forced expiratory volume in the 1st second; IgE, Immunoglobulin-E; FeNO, Fractional Exhaled Nitric Oxide; Severe BSI, moderate-to-severe BSI; SD, standard deviation; IQR, interquartile range; ppb, parts per billion.

|  | Mild BSI<br>(n=7) | Moderate-to-Severe BSI<br>(n=9) | p |
| --- | --- | --- | --- |
| Age, years, mean (SD) | 48 (8.6) | 53.9 (6.8) | 0.24 |
| Male, n (%) | 3 (42.9) | 5 (66.6) | >0.99 |
| BMI, mean (SD) | 23.4 (3.4) | 26.3 (4.6) | 0.37 |
| Duration of asthma, years, mean (SD) | 18.7 (13.2) | 23 (12.1) | 0.48 |
| Positive Skin Prick Test, n (%) | 4 (57.1) | 7 (77.7) | 0.59 |
| CRSwNP, n (%) | 4 (57.1) | 8 (88.8) | 0.26 |
| GERD, n (%) | 2 (28.6) | 1 (18.8) | 0.80 |
| Asthma exacerbations/year, median (IQR) | 6 (3-12) | 12 (4-12) | 0.49 |
| ICS-LABA, n (%) | 16 (100) | 9 (100) | >0.99 |
| LAMA, n (%) | 3 (42.9) | 8 (88.8) | 0.10 |
| Patients with CMH, n (%) | 6 (85.7) | 8 (88.8) | 0.77 |
| Patients on OCS, n, (%) | 3 (42.9) | 7 (77.7) | 0.49 |
| OCS, mg/die, median (IQR) | 25 (0-25) | 25 (1.3-25) | 0.77 |
| ACT, mean (SD) | 16.4 (4.2) | 10.3 (3.5) | **<0.01** |
| FEV<sub>1</sub>, %, mean (SD) | 90.3 (23.5) | 60.8 (17.6) | **<0.01** |
| Blood eosinophil, cells/μL median (IQR) | 718 (500-800) | 640 (370-844) | 0.66 |
| IgE, UI/ml, median (IQR) | 216 (56-431) | 166 (102-144) | 0.46 |
| FeNO, ppb, mean (SD) | 68.3 (63.4) | 32.8 (23.2) | 0.20 |
| Sputum eosinophil, %, median (IQR) | 13 (9-39) | 46 (17.5-66.5) | 0.20 |
| Sputum neutrophil, %, median (IQR) | 27 (10-50) | 22 (6-50) | 0.86 |

**Figure 3.**
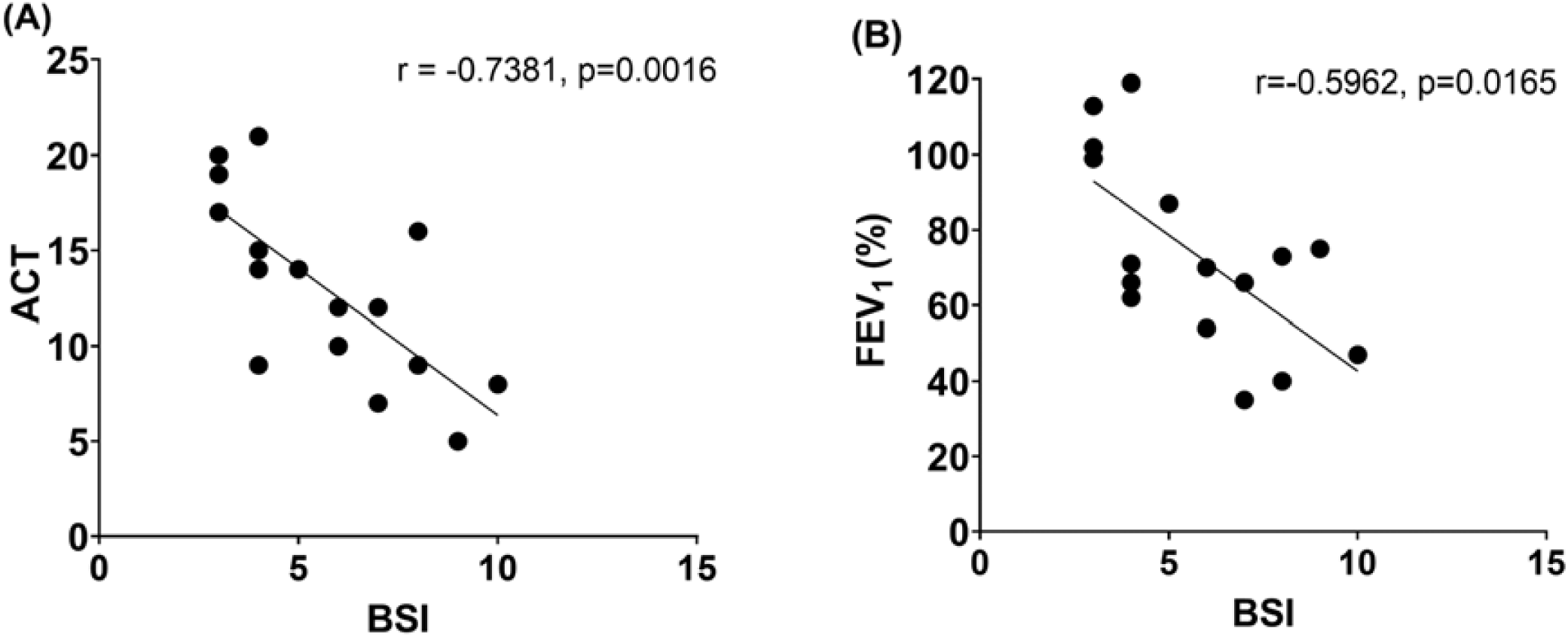
Correlations between BSI, ACT score (panel A) and FEV_1_ (panel B). *Abbreviations: BSI, Bronchiectasis Severity Index; ACT, Asthma Control Test; FEV*_*1*_, *Forced expiratory volume in the 1st second*.

The ACT score was significantly lower in patients with moderate-to-severe BSI compared to the mild BSI group at T0 (10.3±3.5 vs. 16.4±4.2, respectively; p=0.0091). In the mild BSI group, the ACT score increased from 16.4±4.2 to 22.1±4.8 at T12 (p=0.04). In the moderate-to-severe BSI group, the ACT score improved significantly from 10.3±3.5 to 21.2±4 (p=0.0005) and 19.6±6 (p=0.003), at T6 and T12, respectively, as in Figure 4 panel A. Asthma exacerbations/year significantly lowered, from 6 (3-12) to 0 (0-1) in the mild BSI (p=0.015) and from 12 (4-12) to 0 (0-1) in the moderate-to-severe BSI group (p=0.0078), as in Figure 4 panel B. Daily median OCS dose significantly decreased from 25 mg (1.3-25) to 0 (0-0.0) in the moderate-to-severe BSI group (p=0.015), as in Figure 4 panel C. Mean predicted FEV_1_% was significantly lower in the moderate-to-severe BSI group compared to the mild BSI group both at T0 (p=0.008) and T12 (p=0.03). The FEV_1_% significantly increased in the moderate-to-severe BSI group from 60.8±17.6% to 78.2±11.2% at T6 (p=0.01), as shown in Figure 4 panel D. Blood eosinophils count reduced significantly in mild BSI from a median of 718 cells/μL (500-800) to 56 cells/μL (38-120) at T6 (p=0.01). In patients with moderate-to-severe BSI, blood eosinophils decreased from 640 cells/μL (370-844) to 88 cells/μL (42-100) (p=0.007) at T6 and a further reduction to 48 cells/μL (35-102) (p=0.01) was found at T12. Sputum eosinophil count was significantly reduced to a median of 8% (5-23.0) (p=0.015) in mild BSI and 15% (4.5-24.5) (p=0.004) in moderate-to-severe BSI patients, respectively as shown in Figure 5 panel A. Indices of mepolizumab effectiveness, according to BSI, are reported in Table 4.

**Table 4.**
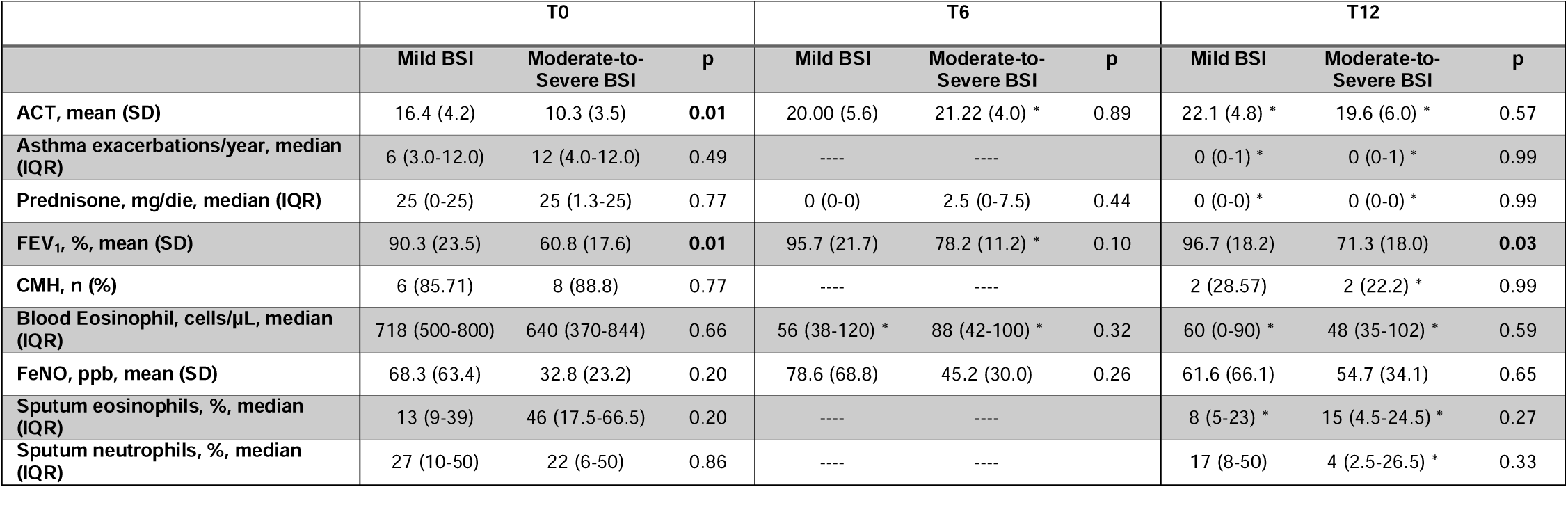
Indices of Mepolizumab effectiveness in severe eosinophilic asthma with associated bronchiectasis (SEA+BE) according to BSI. *p<0.05 T0 vs. T6/T12 *Abbreviations: SEA, Severe Eosinophilic Asthma; BE, Bronchiectasis; BMI, body mass index; CRSwNP, Chronic Rhinosinusitis with Nasal Polyps; GERD, Gastroesophageal reflux disease; ICS-LABA, Inhaled Corticosteroids - Long-Acting Beta-Agonist; LAMA, Long-Acting Muscarinic Antagonist; CMH, Chronic Mucus Hypersecretion; OCS, oral corticosteroids (Prednisone); ACT, Asthma Control Test; FEV*_*1*_, *Forced expiratory volume in the 1st second; IgE, Immunoglobulin-E; FeNO, Fractional Exhaled Nitric Oxide; Severe BSI, moderate-to-severe BSI; SD, standard deviation; IQR, interquartile range; ppb, parts per billion; T0, baseline; T6, follow up at 6 months; T12, follow-up at 12 months*.

**Figure 4.**
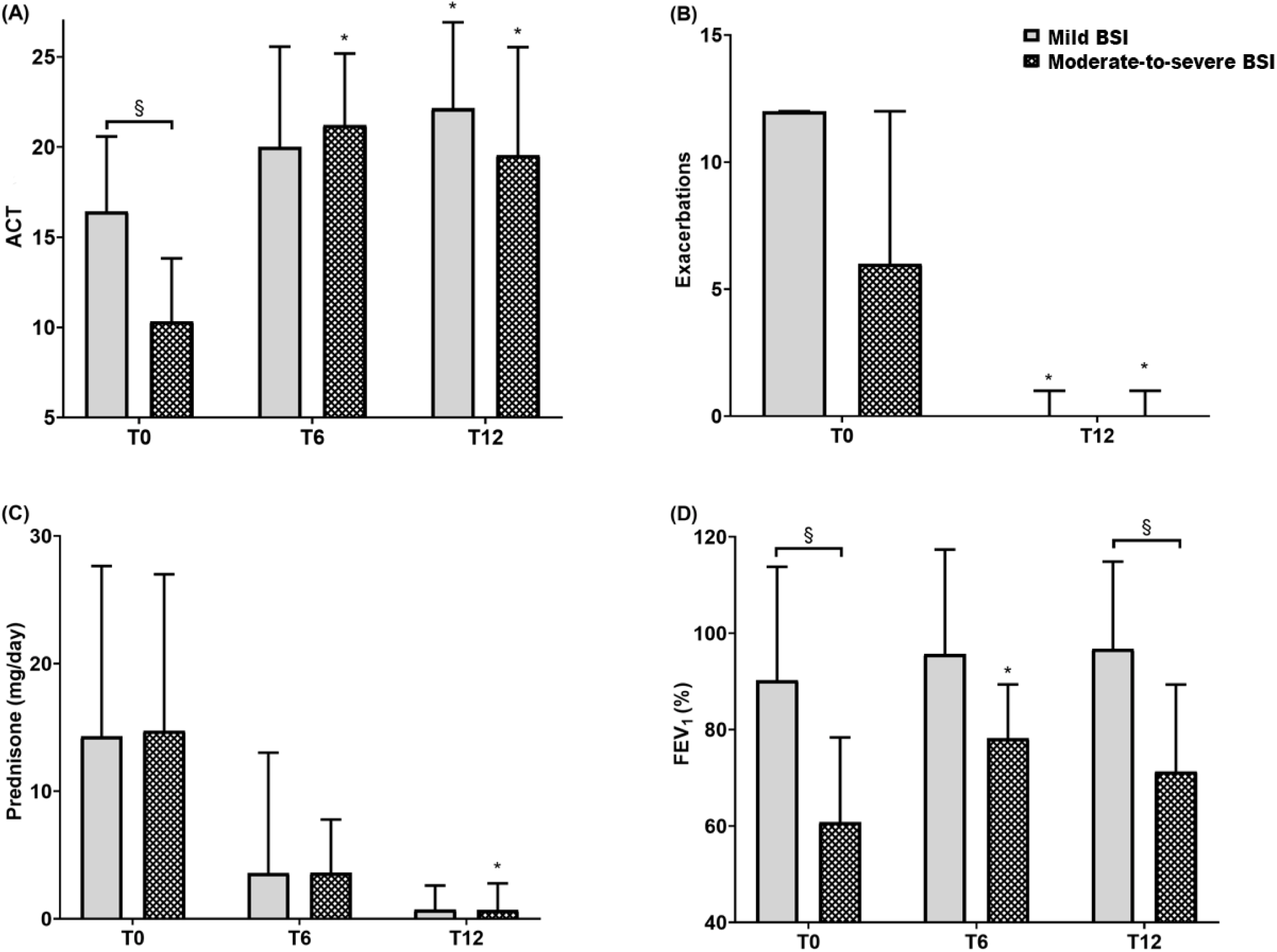
Effects of mepolizumab on ACT score (panel A), asthma exacerbation rate (panel B), OCS intake (panel C), FEV_1_ (panel D) based on the severity of bronchiectasis described as mild BSI and moderate-to-severe BSI groups. ACT score and FEV_1_ values are expressed as mean (SD). All other parameters are expressed as median values (IQR). *p<0.05 T0 vs. T6/T12,§ p<0.05 Mild BSI vs. moderate-to-severe BSI *Abbreviations: BSI, Bronchiectasis Severity Index; ACT, Asthma Control Test; OCS, oral corticosteroids (Prednisone); FEV*_*1*_, *Forced expiratory volume in the 1st second; SD, standard deviation; IQR, interquartile range; ppb, parts per billion, T0, baseline; T6, follow up at 6 months; T12, follow-up at 12 months*.

**Figure 5.**
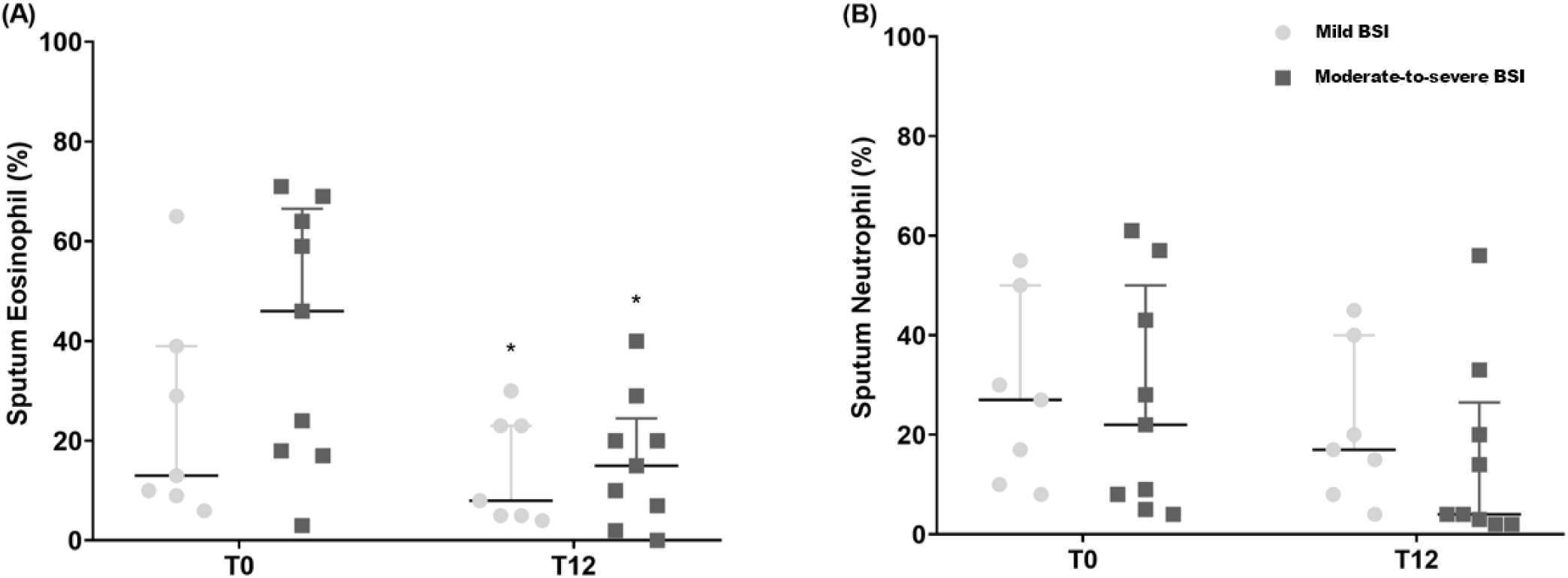
Mepolizumab effects on Sputum Eosinophils % (panel A) and Sputum Neutrophil % (panel B) in Mild BSI and Mild-to-Moderate BSI. *p<0.05 T0 vs. T12 *Abbreviations: BSI, Bronchiectasis Severity Index; SD, standard deviation; IQR, interquartile range; ppb, parts per billion; T0, baseline; T6, follow up at 6 months; T12, follow-up at 12 months*.

## DISCUSSION

To the best of our knowledge, this is the first real-life study assessing the effectiveness of mepolizumab in patients affected by SEA and co-presence of BE in relation to their severity, according to BSI.

Overall, mepolizumab significantly improved asthma symptoms control and lung function and decreased blood eosinophils count, annual exacerbations rate, and OCS intake in the entire population of SEA patients after 6 and 12 months of treatment. These results agree with the published literature on the effectiveness of mepolizumab in this subset of patients [31, 32].

Among our population, we found a high prevalence (50%) of BE in fulfillment with data reported in the literature [5-9]. We also found that patients with SEA+BE showed a significantly higher incidence of chronic mucus hypersecretions than patients with SEA alone; despite this baseline difference, mepolizumab effectively improved chronic mucus secretions in both groups. This represents a clinically significant finding since BE is a muco-obstructive disease characterized by the production of abundant, thick mucus and secretions retention that may worsen asthma symptoms’ perception [33] of patients already at a very severe stage of their disease. Moreover, a clear relationship between mucus hypersecretion and high level of eosinophils in both blood [34] and bronchoalveolar lavage sample [35] has been previously reported, and it might explain our findings.

To date, the way the two diseases affect each other it is still unclear [36]. Of note, in our study, patients with SEA+BE presented a mixed inflammation, with high levels of both eosinophils and neutrophils in sputum samples at baseline that were significantly and concomitantly decreased after treatment. This result further highlights the crucial role of the eosinophilic inflammation in generating bronchiectasis. Thus, eosinophils release granule proteins such as cytotoxic proteins, peroxidases, cationic proteins, and neurotoxins [37,38], determining muco-ciliary epithelium disruption and impairment of muco-ciliary clearance, mucus accumulation and distortion of bronchial architecture. This process might promote chronic bacterial colonization and subsequent neutrophilic airway inflammation [39]. Indeed, elevated eosinophils and neutrophils levels may contribute to stiffening airway mucus gel [40], forming mucus-plugs [41], further worsening airway obstruction [42] and sustaining the Cole vicious circle [40]. Moreover, stimuli such as bacterial and viral infections could cause epithelial cell death and production of IL-25, IL-33, and Thymic Stromal Lymphopoietin that can stimulate Group 2 Innate Lymphoid Cells (ILC-2) proliferation [43] and further production of IL-5 [44].

Mepolizumab, preventing the interaction of IL-5 with its receptor [45], contribute to interrupting this inflammatory pathway, determining the clinical improvement we observed also in the subgroup of patients with SEA+BE. This is a clinically remarkable result since the SEA+BE phenotype is considered refractory to conventional treatment, showing a higher risk of exacerbations, infections, and hospitalization [46]. Our results offer speculation for a tailored treatment approach with mepolizumab of patients with SEA+BE, as already anecdotally reported [8, 47].

We found a statistically significant and clinically meaningful improvement of ACT score in SEA patients with or without co-presence of bronchiectasis, independently of their severity, despite a significantly lower pre-treatment ACT score observed in the most severe patients. From a pulmonary function standpoint, we found a significant improvement in lung function parameters overall after treatment, even in patients with associated bronchiectasis, despite their lower baseline FEV_1_%. Interestingly, we observed a dramatic reduction in the annual rate of asthma exacerbations, which exceed the 50% threshold set up by the British National Institute for Health and Care Excellence (NICE) as a clinically relevant parameter to establish the efficacy of anti-asthma treatments [48]. Furthermore, we found a statistically significant reduction in patients’ daily OCS dose after treatment, in line with data reported by MENSA [49] and SIRIUS [50] trials. Unsurprisingly, eosinophils were dramatically reduced after treatment in blood and sputum samples [51], confirming mepolizumab ability to dampen the cellular responses underlying type 2 asthma [52].

As expected, mepolizumab did not elicit any effects on FeNO levels [53], since NO is stimulated by different pro-inflammatory T2-cytokines, such as IL-4 and IL-13 [54-56].

To the best of our knowledge, our study is the most updated evidence of mepolizumab’s effectiveness in real-world settings, taking into account also complex patients with SEA and co-presence of BE that would have been excluded by clinical trials’ rigid eligibility criteria [57]. These results confirmed previous preliminary data showing the effectiveness of mepolizumab in 4 patients with SEA + BE after three months and one year of treatment [8].

### Strength and limitations

The major strength of the study is the rigorous evaluation of several parameters in all our outpatients with SEA; this allowed us to register the eventual co-presence of BE and assess their severity, using the BSI and the presence of chronic mucus hypersecretion. However, our study has some limitations, mainly related to the relatively small sample size and the retrospective study design.

The limited sample size is mainly influenced by the relatively recent commercial availability and reimbursement of mepolizumab in our geographical area, at the beginning of 2018.

Given the sample size limits and the study design, our further subgroup analysis based on the BSI severity might be underpowered for adequately evaluate treatment effects across subgroups.

## CONCLUSIONS

The results of this study support the hypothesis that mepolizumab elicits similar beneficial effects on ACT and FEV_1_% improvement and exacerbation and daily OCS reduction in patients with SEA with or without coexistence of BE and regardless of their severity. Remarkably, mepolizumab can reduce both eosinophilic and neutrophilic inflammation and chronic mucus secretions, typical SEA+BE phenotype features. This study sets the basis for future research to further support our findings and better understand the role of mepolizumab in treating severe asthma and co-presence of bronchiectasis.

## Data Availability

The datasets generated during and/or analysed during the current study are available from the corresponding author on reasonable request.

## Acknowledgment

none.

## Authors’ contribution

CC conceived the content, drafted the manuscript, approved the final version to be submitted. RI, MP, GC, PI performed data collection, helped in writing the first draft, approved the final version to be submitted. RC, SN, wrote the first draft of the manuscript, performed statistical analysis and approved the final version to be submitted. CP helped in writing the manuscript, revised it critically for important intellectual content and approved the final version to be submitted. NC conceived the content, revised it critically for important intellectual content and approved the final version to be submitted.

## Abbreviations

(SEA): Severe eosinophilic asthma
(BE): Bronchiectasis
(BSI): Bronchiectasis severity index
(BMI): Body mass index
(CRSwNP): Chronic Rhinosinusitis with Nasal Polyps
(GERD): Gastroesophageal reflux disease
(ICS-LABA): Inhaled Corticosteroids - Long- Acting Beta-Agonist
(LAMA): Long-Acting Muscarinic Antagonist
(CMH): Chronic Mucus Hypersecretion
(OCS): oral corticosteroids
(ACT): Asthma Control Test
(FEV1): Forced expiratory volume in the 1st second
(IgE): Immunoglobulin-E
(FeNO): Fractional Exhaled Nitric Oxide
(ERS): European Respiratory Society
(ATS): American Thoracic Society
(GINA): Global Initiative for Asthma.

